# Potential for emergence of foodborne and waterborne trematodiases in California

**DOI:** 10.1101/2022.01.21.22269661

**Authors:** Daniel C.G. Metz, Andrew V. Turner, Alexandria P. Nelson, Ryan F. Hechinger

## Abstract

We document that three human-pathogenic zoonotic trematodes are introduced and widespread throughout southern California in their first intermediate host snail (*Melanoides tuberculata*). We highlight the need to consider these introductions from a public health perspective in California and elsewhere in the United States the snail has invaded.

## Text

Foodborne trematodiases are among the most important neglected infectious diseases of the world, with up towards a billion people estimated to be at risk (Hotez et al. 2008, Keiser & Utzinger 2009, Fürst et al. 2012). These diseases involve a wide range of pathologies, including abdominal pain, chronic cough, hepatomegaly, bile-duct cancer, and brain hemorrhage (Chai et al. 2005, Keiser & Utzinger 2009, Sripa et al. 2010). Foodborne trematodiases result from infection by trematode flatworms that transmit to people who eat second intermediate host organisms carrying infectious metacercariae, which, in turn, originate from larval stages using first intermediate host snails. Foodborne trematodiases have not historically been a major public health concern in the United States, probably given a lack of snails known to transmit injurious trematodes. However, the introduction of one such snail in recent decades opens the door for the emergence of foodborne trematodiases in the United States.

The snail *Melanoides tuberculata* serves as first intermediate host for at least 11 human-infecting trematode species (Pinto & Melo 2011). Although native to Asia and Africa, the snail has been introduced around the world, including in the Americas from the United States to southern Brazil (Murray 1971, Facon et al. 2003, Coelho et al. 2018, CABI 2020, Chalkowski et al. 2021). At least three human-pathogenic, zoonotic trematodes have been co-introduced with the snail in the Americas (see Scholz et al. 2001, Pinto & Melo 2011 and refs therein), including reports in the continental United States of all three species in Texas (Nollen & Murray 1978, Mitchell et al. 2000, Tolley-Jordan & Owen 2008), one in Arizona (Church et al. 2013), and another in Utah and Florida (Mitchell et al. 2005). However, *M. tuberculata* is much more widespread throughout the Unites States (Chalkowski et al. 2021) than the few places the trematodes have been reported. Because these trematodes are dispersed by birds, there is a high probability that the parasites will also be much more widespread in the United States than currently recognized. Despite this possibility, and in contrast to elsewhere in the Americas (e.g., Pulido-Murillo et al. 2018, Lopes et al. 2020), there appears to have been no substantial consideration from a public health perspective of the introduction of *M. tuberculata* and its suite of trematodes in the United States (but see Murray (1971) and Chalkowski et al. (2021)). Here, we document that the snail, its three human-pathogenic trematodes known from elsewhere in the Americas, and several other trematodes potentially introduced to the Americas, are all established in areas of possibly high exposure risk throughout southern California, one of the most populous metropolitan areas in the United States.

In September 2019, we discovered several *M. tuberculata* at a San Diego fishing locality that were infected with *Haplorchis pumilio*, a foodborne human-pathogenic trematode. The finding was surprising because, at that time, not even the snail was reported in the literature as being present in California, nor was it included in the California Department of Fish and Wildlife’s list of invasive invertebrates (https://wildlife.ca.gov/Conservation/Invasives/Species) (but now see Chalkowski et al. 2021). However, observations and data on GBIF.org and iNaturalist.org revealed that *M. tuberculata* was widespread throughout southern California (Figure 1). Given the high probability that the snail’s trematodes would also be widespread, we initiated a broader sampling of southern California. We restricted our focus to freshwater fishing localities, as the most medically important trematodes transmitted by *M. tuberculata* elsewhere in the Americas are fishborne.

**Figure 1.**
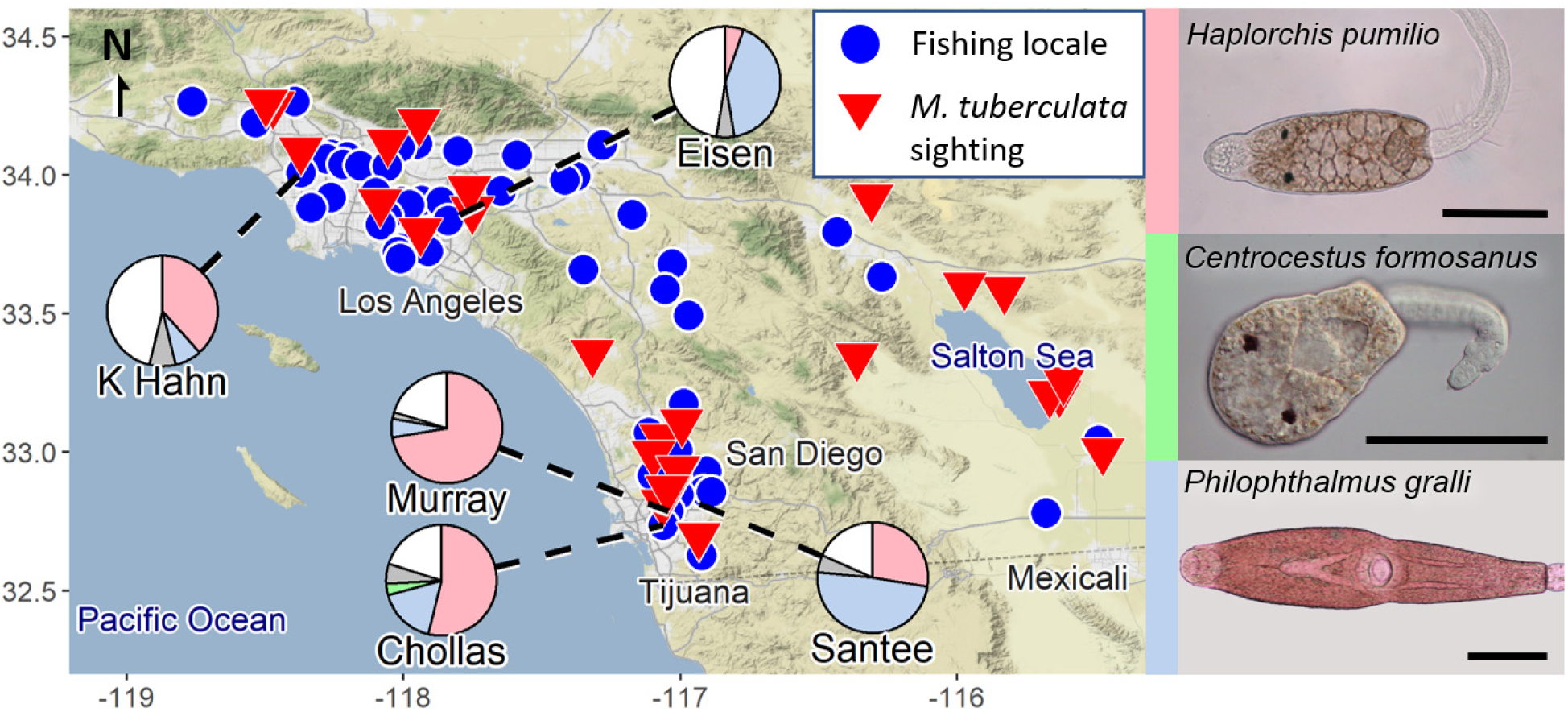
The introduced snail, *Melanoides tuberculata*, and three of its human-pathogenic trematode parasites are widespread throughout southern California. The 25 sites with research-grade *M. tuberculata* observations from iNaturalist.org (bottom points of red inverted triangles) are well-interspersed among the 55 fishing sites (blue circles) listed by the California Department of Fish & Wildlife at elevations likely suitable for *M. tuberculata* survival. Surveyed sites are marked by dashed lines leading from pie charts: K Hahn = Kenneth Hahn State Recreation Area (Los Angeles County), Eisen = Eisenhower Park (Orange Co.), Murray = Murray Reservoir (San Diego Co.), Chollas = Chollas Lake Park (San Diego Co.), Santee = Santee Lakes Recreation Preserve (San Diego Co.). Pie charts indicate parasite prevalence at each surveyed site: green = *Haplorchis pumilio*, gold = *Philophthalmus gralli*, purple = *Centrocestus formosanus*, gray = other non-human-pathogenic trematodes, white = uninfected snails. Photographs of heat-stunned, unstained cercariae of each human-pathogenic trematode are coded to their respective color on the pie charts. Scale bar = 100 μm.

We have thus far hand or dip-net collected live *M. tuberculata* from the shallow waters (<1.2 m) of five southern California fishing localities (Figure 1, Table 1). We dissected snails within 48 hours of collection to identify trematode infections, using microscopical examination to compare worm morphology to literature descriptions (see refs in Pinto & Melo 2011). We extracted genomic DNA from one to five cercariae per infected snail in 1 μL of a 2 mg mL^-1^ proteinase K solution (Qiagen), then amplified 28S rDNA using existing primers (dig12 and 1500R from Tkach et al. (2003)) and PCR protocols (slightly modified from Tkach et al. (2003)). We examined PCR products on a 1x TBE agarose gel before shipping successful products to Eton Biosciences (San Diego, CA) for Sanger sequencing services. We used MEGA X (Stecher et al. 2020) to trim the trace files by eye prior to a discontiguous megaBLAST comparing each sequence to all 28S Platyhelminthes sequences present in GenBank (http://www.ncbi.nlm.nih.gov) as of September 2021. We archived newly generated sequences in GenBank (Table 1).

**Table 1.** Prevalences (±95% CI *) of human-infecting and other trematode species in the introduced first intermediate host snail *Melanoides tuberculata* from our initial sample sites in southern California, USA.

| Parasite | Locality<sup>†</sup> |  |  |  |  |  |
| --- | --- | --- | --- | --- | --- | --- |
|  | San Diego Co. |  |  | Orange Co. | Los Angeles Co. | Average among sites (weighted by n) |
|  | Murray Reservoir | Santee Lakes | Chollas Lake | Eisenhower Park Lake | Kenneth Hahn SRA Lake |  |
| *Haplorchis pumilio*<sup>‡</sup> | 72.2 (59.1-82.4) | 27.8 (19.9-37.5) | 55.4 (49.7-61) | 5.3 (0.3-24.6) | 38.5 (17.7-64.5) | 49.3 |
| *Philophthalmus gralli*<sup>‡</sup> | 5.6 (1.9-15.1) | 49.5 (39.7-59.3) | 17.2 (13.4-21.9) | 42.1 (23.1-63.7) | 7.7 (0.4-33.3) | 23.2 |
| *Centrocestus formosanus*<sup>‡</sup> | 0 (0-6.6) | 0 (0-3.8) | 3.4 (1.8-6.1) | 0 (0-16.8) | 0 (0-22.8) | 2.1 |
| Renicolid sp. 1<sup>§</sup> | 0 (0-6.6) | 4.1 (1.6-10.1) | 4.7 (2.8-7.8) | 0 (0-16.8) | 0 (0-22.8) | 3.7 |
| Renicolid sp. 2<sup>¶</sup> | 0 (0-6.6) | 0 (0-3.8) | 0.7 (0.2-2.4) | 0 (0-16.8) | 0 (0-22.8) | 0.4 |
| Renicolid sp. 3<sup>#</sup> | 0 (0-6.6) | 1 (0.1-5.6) | 0 (0-1.3) | 0 (0-16.8) | 0 (0-22.8) | 0.2 |
| Lecithodendriid gen. sp.** | 1.9 (0.1-9.8) | 0 (0-3.8) | 0.3 (0-1.9) | 5.3 (0.3-24.6) | 7.7 (0.4-33.3) | 0.8 |
| Plagiorchid? sp. 1<sup>††</sup> | 0 (0-6.6) | 0 (0-3.8) | 0.3 (0-1.9) | 0 (0-16.8) | 0 (0-22.8) | 0.2 |
| Uninfected | 20.4 (11.8-32.9) | 18.6 (12.1-27.4) | 20.6 (16.4-25.6) | 47.4 (27.3-68.3) | 46.2 (23.2-70.9) | 21.9 |
| Total no. snails<sup>‡‡</sup> | 54 | 97 | 296 | 19 | 13 | 479 |

*M. tuberculata* harbored introduced trematode infections at each of the five localities. Two trematode species morphologically and genetically (99.8-100%) matched the fishborne trematodes *Haplorchis pumilio* and *Centrocestus formosanus*, which are introduced elsewhere in the Americas (Mitchell et al. 2000, Scholz & Salgado-Maldonado 2000, Tolley-Jordan & Owen 2008, Pulido-Murillo et al. 2018, Lopes et al. 2020). *H. pumilio* was the most common trematode, being present at each locality at relatively high prevalences (Table 1, Figure 1). These introduced heterophyids have garnered attention given the impacts on second intermediate host fish (e.g., Mitchell et al. 2000, Mitchell et al. 2005, Huston et al. 2014). However, like other heterophyids, the adult stages of these trematodes cause a wide range of pathologies in humans, sometimes possibly being deadly (Keiser & Utzinger 2009, Sripa et al. 2010, Chai & Jung 2017). People are infected when eating raw, undercooked, or pickled second intermediate host fish, or via utensil contamination. Because these parasites occur in fishing localities, can infect a wide range of fishes, and fish are commonly eaten in California, including uncooked as ceviche, poke, and sushi (unpub. obs.), there is a real possibility for *H. pumilio* and *C. formosanus* to cause foodborne trematodiasis in California and elsewhere in the United States.

The other encountered human-pathogenic trematode morphologically and genetically (99.6-100%) matched *Philophthalmus gralli*, which is also reported from *M. tuberculata* in Arizona (Church et al. 2013), Texas (Nollen & Murray 1978) and farther south in the Americas (Diaz et al. 2002, Chalkowski et al. 2021). *P. gralli* infects humans via ingestion of aquatic hosts harboring metacercariae or by direct contact with swimming infectious cercariae. *Philophthalmus* species can infect the eyes of humans and cause conjunctivitis (Gutierrez et al. 1987), with a recent case in Texas (Sapp et al. 2019). *P. gralli* can also cause substantial problems in captive birds (e.g., Greve & Harrison 1980, Church et al. 2013). Although *P. gralli* may be of less human health concern than the above two species, it was the second most common trematode (Table 1) among the three introduced human-infecting trematodes transmitted by *M. tuberculata* in California and elsewhere in the United States.

We detected five additional trematode species, including two not previously reported from *M. tuberculata* in the Americas, all of which are possibly introduced (Table 1). These species are likely of little direct public health concern given their probable life cycles, but they may be of veterinary interest (Table 1). However, if they do represent novel introductions to the Americas, their presence highlights that additional foodborne trematodes transmitted by *M. tuberculata* in its native range may also be introduced here. Such trematodes include severely pathogenic trematodes such as the carcinogenic human liver flukes (*Clonorchis sinensis* and *Opisthorchis* species), and the lung fluke (*Paragonimus westermani*).

Hence, several factors suggest the possible emergence and even ongoing presence of foodborne trematodiases in California: (1) the introduced *M. tuberculata* snail is widespread, including at localities where people catch fish for eating; (2) the snail carries at least three of its human-pathogenic trematodes, including two that are fishborne; and (3) the snail also carries several other potentially introduced trematodes, indicating the possibility that additional human-pathogenic trematodes are already introduced or may be introduced in the future. Taken together, these factors clearly call for additional consideration of *M. tuberculata* from a public and veterinary health perspective in California and wherever else the snail is present in the United States.

## Data Availability

All data produced in the present work are contained in the manuscript.

## Acknowledgements

We thank Emma Palmer, Jordan Ingco and Taylor Ackerknecht for field/laboratory assistance. This work was supported by the US National Institutes of Health Grant# 1R03AI156569-01.

## References

CABI (2020) Datasheet: Melanoides tuberculata (red-rimmed melania) [original text by B Facon and JP Pointier]. Invasive Species Compendium. CAB International, Wallingford, UK

Chai J-Y, Darwin Murrell K, Lymbery AJ (2005) Fish-borne parasitic zoonoses: Status and issues. Int J Parasitol 35:1233–1254

Chai J-Y, Jung B-K (2017) Fishborne zoonotic heterophyid infections: An update. Food and Waterborne Parasitology 8-9:33–63

Chalkowski K, Abigail M, Christopher AL, Sarah Z (2021) Spread of an Avian Eye Fluke, Philophthalmus gralli, through Biological Invasion of an Intermediate Host. J Parasitol 107:336–348

Church ML, Barrett PM, Swenson J, Kinsella JM, Tkach VV (2013) Outbreak of Philophthalmus gralli in four greater rheas (Rhea americana). Vet Ophthalmol 16:65–72

Coelho PN, Fernandez MA, Cesar DAS, Ruocco AMC, Henry R (2018) Updated distribution and range expansion of the gastropod invader Melanoides tuberculata (Muller, 1774) in Brazilian waters. Bioinvasions Records 7:405–409

Diaz MT, Hernandez LE, Bashirullah AK (2002) Experimental life cycle of Philophthalmus gralli (Trematoda : Philophthalmidae) in Venezuela. Rev Biol Trop 50:629–641

Facon B, Pointier JP, Glaubrecht M, Poux C, Jarne P, David P (2003) A molecular phylogeography approach to biological invasions of the New World by parthenogenetic Thiarid snails. Mol Ecol 12:3027–3039

Fürst T, Keiser J, Utzinger J (2012) Global burden of human food-borne trematodiasis: a systematic review and meta-analysis. The Lancet Infectious Diseases 12:210–221

Greve JH, Harrison GJ (1980) Conjunctivitis caused by eye flukes in captive-reared ostriches. J Am Vet Med Assoc 177:909–910

Gutierrez Y, Grossniklaus HE, Annable WL (1987) Human conjunctivitis caused by the bird parasite Philophthalmus. American Journal of Ophthalmology 104:417–419

Hotez PJ, Brindley PJ, Bethony JM, King CH, Pearce EJ, Jacobson J (2008) Helminth infections: the great neglected tropical diseases. Journal of Clinical Investigation 118:1311–1321

Huston DC, Worsham MD, Huffman DG, Ostrand KG (2014) Infection of fishes, including threatened and endangered species by the trematode parasite Haplorchis pumilio (Looss, 1896) (Trematoda: Heterophyidae). Bioinvasions Records 3:189–194

Keiser J, Utzinger J (2009) Food-Borne Trematodiases. Clinical Microbiology Reviews 22:466–483

Lopes AS, Pulido-Murillo EA, López-Hernández D, Melo ALd, Pinto HA (2021) First report of Melanoides tuberculata (Mollusca: Thiaridae) harboring a xiphidiocercaria in Brazil: A new parasite introduced in the Americas? Parasitol Int 82:102284

Lopes AS, Pulido-Murillo EA, Melo AL, Pinto HA (2020) Haplorchis pumilio (Trematoda: Heterophyidae) as a new fish-borne zoonotic agent transmitted by Melanoides tuberculata (Mollusca: Thiaridae) in Brazil: A morphological and molecular study. Infection, genetics and evolution : journal of molecular epidemiology and evolutionary genetics in infectious diseases 85:104495–104495

Mitchell AJ, Overstreet RM, Goodwin AE, Brandt TM (2005) Spread of an exotic fish-gill trematode. Fisheries 30:11–16

Mitchell AJ, Salmon MJ, Huffman DG, Goodwin AE, Brandt TM (2000) Prevalence and pathogenicity of a heterophyid trematode infecting the gills of an endangered fish, the fountain darter, in two central Texas spring-fed rivers. J Aquat Anim Health 12:283–289

Murray HD (1971) The introduction and spread of thiarids in the USA. Biologist (Charlest) 53:133–135

Nollen PM, Murray HD (1978) Philophthalmus gralli - identification, growth characteristics, and treatment of an oriental eyefluke of birds introduced into continental United States. J Parasitol 64:178–180

Pinto HA, Melo AL (2011) A checklist of trematodes (Platyhelminthes) transmitted by Melanoides tuberculata (Mollusca: Thiaridae). Zootaxa:15–28

Pinto HA, Melo AL (2012) Melanoides tuberculata (Mollusca: Thiaridae) harboring renicolid cercariae (Trematoda: Renicolidae) in Brazil. J Parasitol 98:784–787

Pulido-Murillo EA, Furtado LFV, Melo AL, Rabelo EML, Pinto HA (2018) Fishborne Zoonotic Trematodes Transmitted by Melanoides tuberculata Snails, Peru. Emerg Infect Dis 24:606–608

Sapp SGH, Alhabshan RN, Bishop HS, Fox M, Ndubuisi M, Snider CE, Bradbury RS (2019) Ocular Trematodiasis Caused by the Avian Eye Fluke Philophthalmus in Southern Texas. Open Forum Infectious Diseases 6

Scholz T, Aguirre-Macedo ML, Salgado-Maldonado G (2001) Trematodes of the family Heterophyidae (Digenea) in Mexico: a review of species and new host and geographical records. J Nat Hist 35:1733–1772

Scholz T, Salgado-Maldonado G (2000) The Introduction and Dispersal of Centrocestus formosanus (Nishigori, 1924) (Digenea: Heterophyidae) in Mexico: A Review. The American Midland Naturalist 143:185–200

Sripa B, Kaewkes S, Intapan PM, Maleewong W, Brindley PJ (2010) Food-Borne Trematodiases in Southeast Asia: Epidemiology, Pathology, Clinical Manifestation and Control. In: Zhou XN, Bergquist R, Olveda R, Utzinger J (eds) Advances in Parasitology, Vol 72: Important Helminth Infections in Southeast Asia: Diversity and Potential for Control and Elimination, Pt A, Book 72

Stecher G, Tamura K, Kumar S (2020) Molecular Evolutionary Genetics Analysis (MEGA) for macOS. Mol Biol Evol 37:1237–1239

Tkach VV, Littlewood DTJ, Olson PD, Kinsella JM, Swiderski Z (2003) Molecular phylogenetic analysis of the Microphalloidea Ward, 1901 (Trematoda: Digenea). Syst Parasitol 56:1–15

Tolley-Jordan LR, Owen JM (2008) Habitat influences snail community structure and trematode infection levels in a spring-fed river, Texas, USA. Hydrobiologia 600:29–40

